# When to relax a lockdown? A modelling-based study of testing-led strategies coupled with sero-surveillance against SARS-CoV-2 infection in India

**DOI:** 10.1101/2020.05.29.20117010

**Authors:** Sandip Mandal, Hemanshu Das, Sarang Deo, Nimalan Arinaminpathy

## Abstract

India’s lockdown against SARS-CoV-2, if lifted without any other mitigations in place, could risk a second wave of infection. A test-and-isolate strategy, using PCR diagnostic tests, could help to minimise the impact of this second wave. Meanwhile, population-level serological surveillance can provide valuable insights into the level of immunity in the population. Using a mathematical model, consistent with an Indian megacity, we examined how seroprevalence data could guide a test-and-isolate strategy, for lifting a lockdown. For example, if seroprevalence is 20% of the population, we show that a testing strategy needs to identify symptomatic cases within 5 – 8 days of symptom onset, in order to prevent a resurgent wave from overwhelming hospital capacity in the city. This estimate is robust to uncertainty in the effectiveness of the lockdown, as well as in immune protection against reinfection. To set these results in their economic context, we estimate that the weekly cost of such a PCR-based testing programme would be less than 2.1% of the weekly economic loss due to the lockdown. Our results illustrate how PCR-based testing and serological surveillance can be combined to design evidence-based policies, for lifting lockdowns in Indian cities and elsewhere.

## Introduction

The emergence of the novel virus SARS-CoV-2 has prompted stringent physical distancing measures around the world. In India, one of the world’s most populous countries, a countrywide ‘lockdown’ has been in effect since March 25^th^ 2020^1^. The lockdown involves blanket restrictions on movement, closure of shops with the exception of essential services, closure of all school and universities, and banning of social gatherings^2^. Similar measures in China and elsewhere have succeeded in slowing transmission^3,4^. However, such measures also inflict severe societal and economic disruption. Consequently, there is increasing attention in India – and in many other countries under similar restrictions – on how best to relax such control measures, while preserving public health imperatives^5^.

Previous work has illustrated that the relaxing of a lockdown, if conducted too rapidly, could risk another surge of infection, that can overwhelm health systems^6,7^. Some proposed strategies include (i) a cycle of lockdowns and releases^6,8^, to maintain any resurgent epidemic to within levels manageable by the health system, and (ii) the use of serological tests to identify those who have had exposure to the virus, and might thus be presumed safe to return to normal activity^9^. For the latter to be implemented, there needs to be high confidence in the serological test being used, particularly that is has minimal risk of false positivity. The former strategy may face real challenges in such a complex society as in India, particularly in view of the effect of such measures amongst the most disadvantaged^10^.

Another strategy involves the use of systematic and intensive testing, to identify and quarantine cases of SARS-CoV-2 infection as early as possible. This approach poses real logistical challenges, not least the need to establish a population-wide system of contact tracing, together with reliable, readily accessible network of testing facilities with ample capacity, providing accurate monitoring and support for those who are diagnosed and isolated. Such approaches are only feasible when prevalence is low and infections are clustered in nature; they have been implemented successfully in South Korea, Taiwan and elsewhere^11,12^, to slow the initial pandemic waves and to allow health systems to cope.

For countries that instead had to impose population-wide lockdowns, community-based testing programmes could still be invaluable, as part of a strategy to allow the lockdown to be lifted, while still protecting the population. Essentially, testing offers a means for moving from a blanket lockdown to targeted quarantine, more so for urban areas with high transmission probability in densely populated pockets. However, such an approach raises important questions of its own. For example, how can health authorities assess whether cases in the community are being identified and isolated sufficiently rapidly, to safely lift the lockdown? In low- and middle-income settings, where surveillance data is sparse, there is real uncertainty about the degree of transmission that had already occurred by the time of the lockdown, as well as the impact that the lockdown itself is having on transmission. How should decisions about post-lockdown testing strategy be made in the face of such uncertainty?

Here we address these questions using a dynamical mathematical model of SARS-CoV-2 transmission. Using this model, we illustrate the value of setting up a two-pronged testing capacity for both surveillance (using serological testing) and outbreak control. Appropriately coordinated, surveillance could provide valuable strategic information on the intensity of community-based testing that would be needed in order to allow the lockdown to be lifted. To place these results in their economic context, we provide preliminary estimates for the economic impact of a lockdown and compare it with the cost of testing. Although our analysis focuses on Indian metropolitan areas, it offers insights for other, similar settings that face uncertainties about when a lockdown can safely be lifted.

## Methods

The model is a compartmental, deterministic framework, described here in outline, and with further technical details given in the appendix. For the purpose of illustration, we focus on a megacity akin to India. To take account of the highly age-specific patterns of severity of SARS-nCoV-2 infection^13^, as well as age demographics typical of megacities in India, the model is stratified into three age groups: < 15 years old, 16 – 64 years old, and > 65 years old.

There is broad evidence that asymptomatic infection can contribute to transmission^14,15^. The model distinguishes asymptomatic vs presymptomatic infection, with parameters drawn from previous analysis on the relative infectiousness of both^6^. In the absence of robust estimates for the basic reproduction number, R0 in India, we assume a range of scenarios from 2 to 3.

### Interventions

We modelled the effects of a lockdown in a simple way, by assuming that it reduces transmission across the whole population by a given amount, denoted as its ‘effectiveness’. We assumed that the lockdown is lifted when hospital bed occupancy due to COVID-19 declines to 10% of total bed availability in the megacity. (The specific choice of trigger is not critical for the purpose of the current study.) At this point we assumed that the lockdown is gradually lifted over two weeks, with the overall community contact rate being restored to its full value over this time.

We denote the ‘impact’ of a lockdown as the reduction that it achieves, in overall illness and mortality due to COVID-19, and the ‘effectiveness’ of a lockdown as the daily reduction in transmission intensity that it causes. Impact depends not only effectiveness, but also on the timing of lockdown initiation with respect to the phase of epidemic. As illustrated in Figure S1 in the appendix, a lockdown that is initiated late in an epidemic can have a markedly worse outcome than an equally strong lockdown that is initiated early. However, the available surveillance data does not allow a robust estimate of either effectiveness nor timing. For the purpose of illustrative simulations, we adopted scenarios with given assumptions for both parameters. For subsequent, analytical simulations, we then sampled from a range of scenarios for the effectiveness and timing of the lockdown, as well as for the role of asymptomatic infection in transmission, and for the role of immunity in protection against reinfection (see Table 1, and Table S1 in the appendix).

**Table 1.** Additional sources of uncertainty used in Figures 3 and 4. See Table S1 for further parameter values.

| Parameter | Range | Notes |
| --- | --- | --- |
| *Lockdown* |  |  |
| Timing of lockdown, relative to epidemic | At time of $n$-th COVID-related death, where $n$ is between 1 and 1,000 | Lockdown timing can have strong influence on its effect (see Fig.S2, appendix). Here we adopt a wide range of possibilities for numbers of actual deaths before lockdown. |
| Effectiveness of lockdown in reducing transmission | 15 – 85% | Lockdown effectiveness depends on compliance, and on achievable physical distancing in crowded settings. Again adopting wide uncertainty ranges given lack of systematic data |
| *Asymptomatic transmission* |  |  |
| Proportion of cases being asymptomatic throughout their infection | 10 – 33% | Parameters relating to the role of asymptomatic infections, for transmission. Scenarios having a strong role for such infections would diminish the impact of isolating symptomatic infections alone. |
| Relative infectiousness of asymptomatic and presymptomatics vs symptomatic infection | 10 – 66% |  |
| *Post-infection immunity* |  |  |
| Average duration of immunity | > 6 months | Studies with established human coronaviruses suggest immunity lasting at least a year <sup>32</sup>; for SARS-CoV-2 we assume a more pessimistic scenario of at least 6 months |
| Susceptibility to reinfection, relative to susceptibility to primary infection | 0 – 10% | 0% indicates that immunity is completely protective to reinfection |

We simulated two types of scenarios for the lifting of a lockdown: first, that the lockdown is relaxed with no other interventions in place, and second, that an intensive testing programme is established two weeks before the lockdown is lifted, and maintained indefinitely thereafter. For the latter scenario, we assumed a two-pronged testing programme that utilises: (i) real-time RT-PCR tests to identify and isolate infectious cases in the community, along with (ii) laboratory-based sero-surveillance in a population-representative group, in order to monitor the development of population immunity over time. We modelled the test-and-isolation activities in a simple way, assuming that symptomatic individuals are identified and successfully isolated, within a given average delay from symptom onset.

### Cost of testing vs cost of lockdown

To set these findings in an economic context, we attempted to estimate the economic loss arising from each week of lockdown in an Indian megacity similar to Delhi, and to compare it with the potential cost of a community-level RT-PCR-testing programme. For economic loss, we based our estimates on the loss in productivity due to shutdown of activities, separately for each sector. We then combined the productivity loss in each sector with the sectoral composition, taking the state of Delhi as an illustrative example, to generate the overall economic loss per week of lockdown. For the cost of a testing programme, we considered an illustrative scenario, wherein all COVID-19 symptomatics in the megacity are tested sufficiently frequently to be identified and isolated within 5 days of symptom onset. As discussed below, this scenario is artificially resource-intensive, but serves to provide an upper bound on the potential cost of a testing programme. For this scenario we adopted an activity-based costing approach, which covered sample collection and transportation along with sample testing. Further details are provided in the supporting information, under sections 2 and 3.

## Results

Figure 1A illustrates model projections for the numbers of people needing hospitalisation with SARS-CoV-2, in a megacity. The figure illustrates different scenarios for the strength of the lockdown (ranging from a 25% reduction of transmission to 75%), as well as the potential implications of ultimately lifting the lockdown. In the event that the lockdown is effective in controlling transmission (yellow curve), its release results in a resurgent epidemic, that can be sufficiently severe to overwhelm the health system, i.e. with the number of persons needing hospitalization exceeding the existing hospital capacity. However, a less effective lockdown (red curve) does not see any resurgent epidemic upon being lifted.

**Figure 1.**
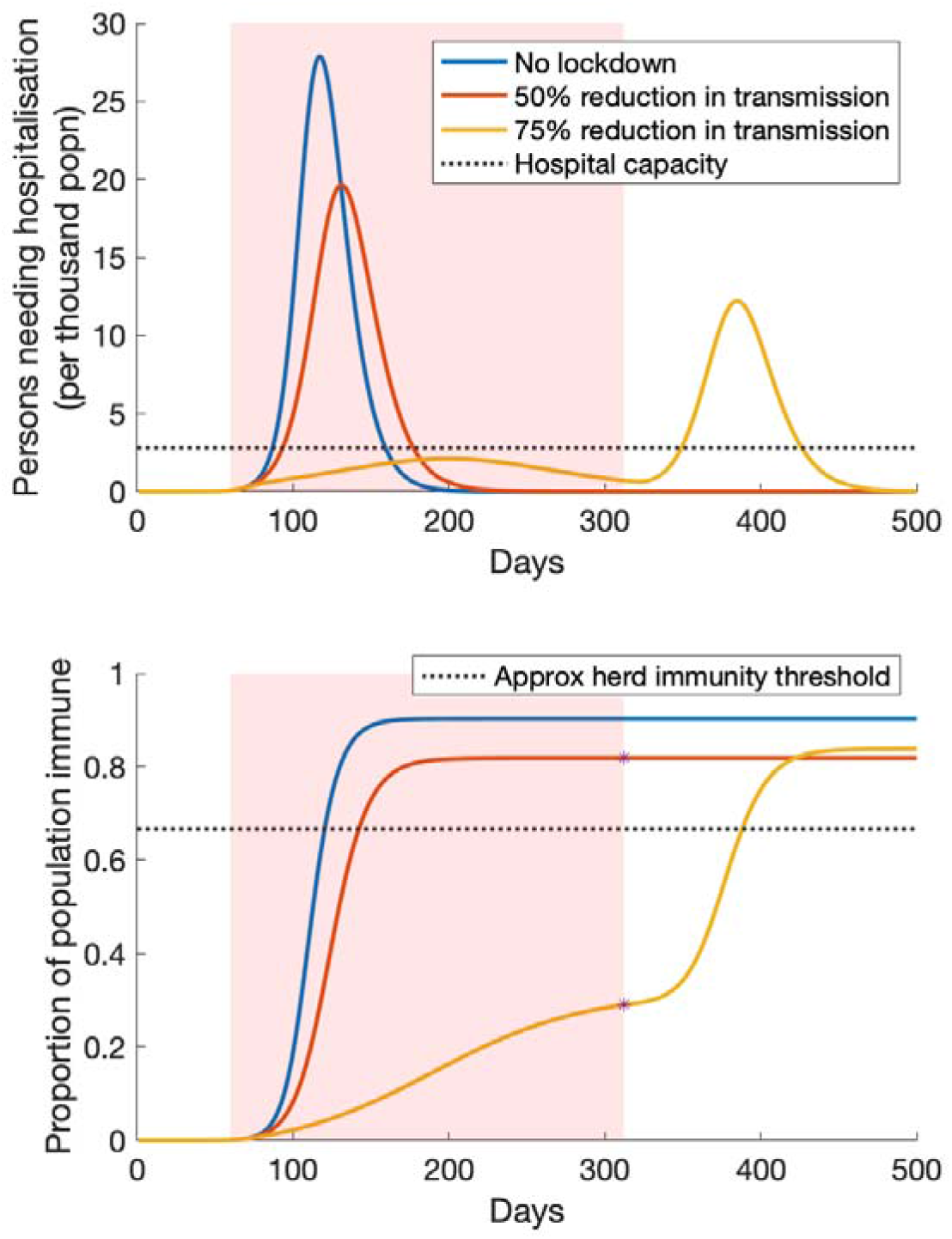
Illustration of lockdown release, with no other interventions in effect. Shown are three scenarios: no lockdown (blue curve); an ineffective lockdown that reduces transmission by only 25% (red curve); and a more effective lockdown that reduces transmission by 75% (yellow) curve. The pink-shaded area denotes the duration of the lockdown: we assume that it is lifted when COVID-related hospitalisations decline to 10% of capacity. Upper panel: numbers of individuals needing hospitalisation over time. Lower panel: the accumulation of population immunity over time, for the scenarios simulated in the upper panel. Stars on the red and yellow curves indicate the level of population immunity that would be estimated by sero-surveillance, at the time of lifting the lockdown. The dashed line indicates the approximate level of herd immunity (approximate because it is calculated as 1 1/, a formula for homogenous populations): a population having a higher level of immunity would effectively be protected from a resurgent epidemic, without need for further interventions or lockdown.

We note that these overall dynamics are consistent with previous modelling findings in other settings^6^. Figure 1B helps to explain these dynamics, in terms of population immunity. A poorly effective lockdown allows sufficient infection in the population, to exceed the herd immunity threshold, i.e, the level of population immunity at which the virus can no longer sustain transmission. By contrast a highly effective lockdown, through limiting transmission, creates a population that has not achieved herd immunity: that is, a population that is vulnerable to a resurgent epidemic, upon lifting of the lockdown.

Assuming a population under an effective lockdown, we next examine how such a population might be protected through an intensive test-and-isolate strategy, that accompanies the lifting of the lockdown. Figure 2A illustrates the potential implications of such a strategy, assuming that it is instated two weeks prior to the lifting the lockdown, and is sufficiently intensive to isolate all symptomatic individuals within, on average, 4 days of symptom onset. Even though a resurgent epidemic occurs, the figure illustrates that it is within levels that do not overwhelm the health system. Thus the lockdown, not only impacts transmission, but also performs the critical role of allowing time to build up the necessary testing capacity, to ultimately permit the lockdown safely to be lifted. Figure 2B again shows the accumulation of population immunity under these different scenarios.

**Figure 2.**
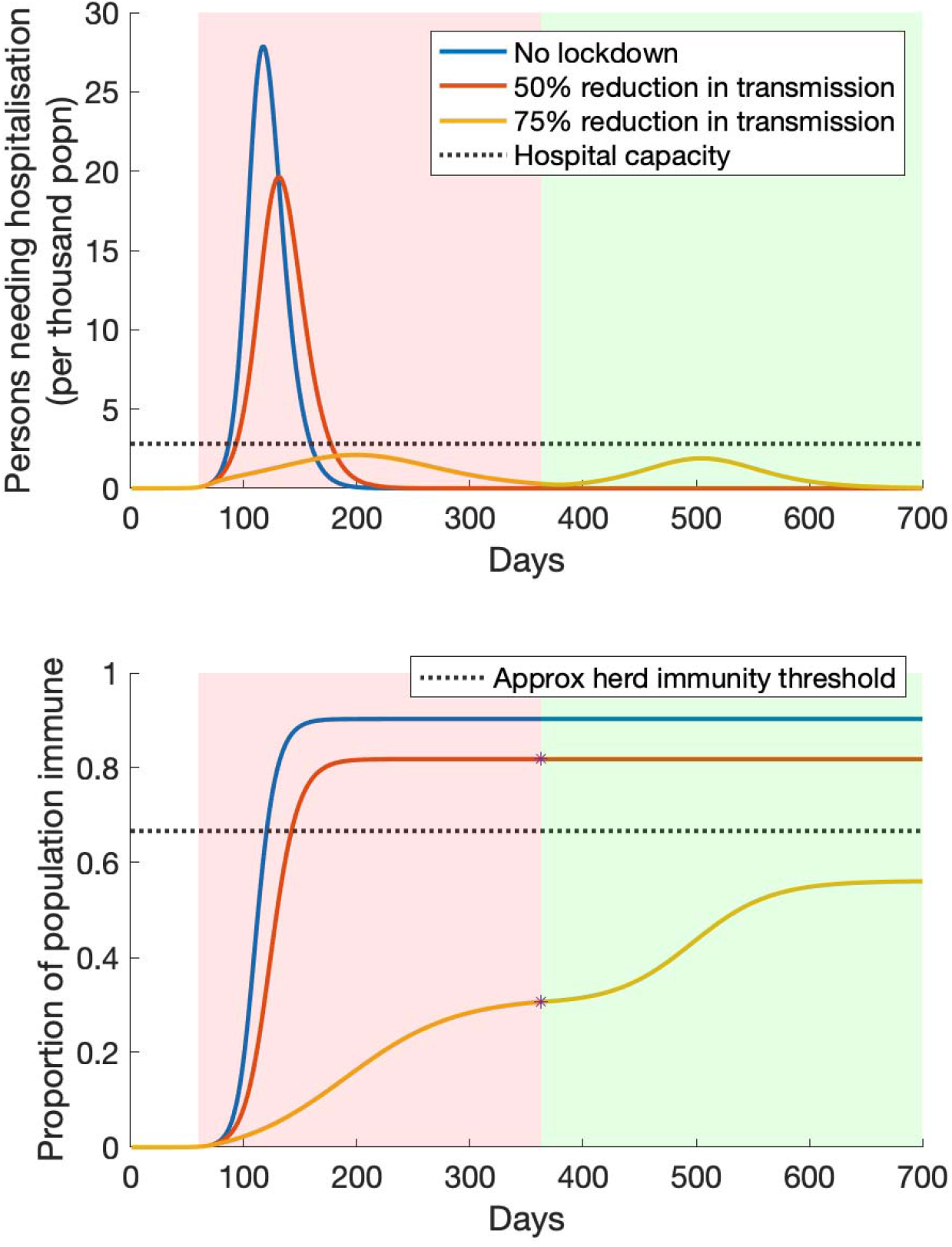
Illustration of lockdown release, when accompanied by a test-and-isolate strategy. Scenarios are as in Figure 1, but here assuming that – two weeks prior to the lifting of the lockdown – PCR testing in the community is sufficiently ramped up to identify and isolate all symptomatics within, on average, 4 days on symptom onset. As in Figure 1, the pink shaded area denotes the duration of the lockdown; the green shaded area denotes the period when the lockdown is lifted, and the test-and-isolate strategy is implemented in its place. In the event that the lockdown has been effective in reducing transmission (yellow curve), such a strategy succeeds in preventing the resurgent epidemic from overwhelming healthcare capacity. In the event that the lockdown has been less effective (red curve), such a strategy has little additional value for epidemic control, since population immunity is already sufficiently widespread to interrupt transmission (lower panel).

These figures illustrate how sero-surveillance could be invaluable in the face of uncertainty, including about the effectiveness of a lockdown. In Figure 1, for example, estimates of seroprevalence can help to establish whether the risk of a resurgent epidemic is low (as on the red curve) or high (as on the yellow curve), at the point of lifting a lockdown. Therefore, in Figure 2, it is reasonable to hypothesise that such data can help inform the intensity of testing required, in order for a lockdown to be safely lifted, i.e. without overwhelming the health system. For example, if population immunity is known, then how rapidly should symptomatic cases be diagnosed and isolated? Is it adequate to test symptomatic cases only, or is there a need to identify and isolate asymptomatic cases as well?

Figure 3 addresses these questions, for a virus having *R*_0_ = 2.5. For this figure we now allow wide uncertainty in the lockdown effectiveness, and in its timing with respect to the epidemic. Each point in Figure 3A represents a distinct scenario for each of these unknown parameters, as well as for the importance of asymptomatic infection in transmission, and for the role of immunity in protecting against reinfections (see table 1 for details). For each scenario, we simulate a lockdown that is released when COVID-related hospitalisations ultimately decline to 10% of the hospital bed capacity. We then record the proportion of the population immune at this point in the epidemic (x-axis of Figure 3A), as well as the minimum intensity of symptomatic testing that is required, in order to prevent a resurgent epidemic from overwhelming the health system (y-axis of Figure 3A). The figure illustrates that – despite the various uncertainties incorporated in these projections – there remains a coherent relationship between population immunity, and the intensity of testing that is required to protect a population from a resurgent epidemic. For instance, if the seroprevalence in the population at the point of lifting the lockdown is 20%, Figure 3 suggests that a resurgent epidemic could be prevented from overwhelming the hospital bed capacity as long as symptomatic cases are identified and isolated within 5 – 8 days of symptom onset (vertical spread of points along the y-axis at 20% seroprevalence).

**Figure 3.**
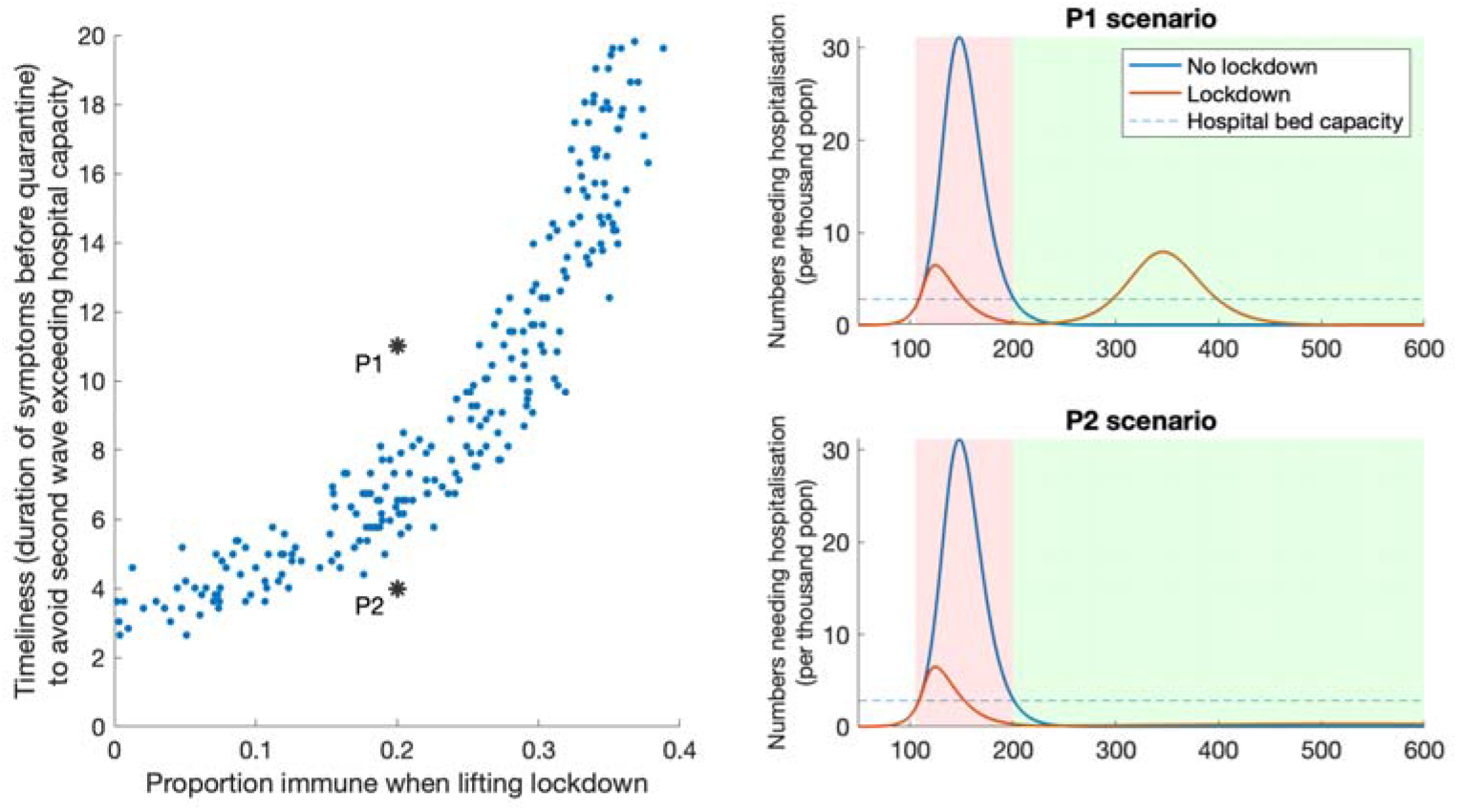
How sero-surveillance can guide post-lockdown strategy, with R0 = 2.5. Left hand panel shows how sero-surveillance (x-axis) can guide decisions about the required intensity of post-lockdown PCR testing in the community (y-axis). The figure is divided into two zones, by the ‘band’ of points. In the lower zone, population immunity is high enough, and the PCR testing strategy is intense enough, to prevent a resurgent epidemic from overwhelming the health system – and conversely in the upper zone. Points P1 and P2 show examples in both zones, their dynamics illustrated in the right-hand panels. As in Figure 2, pink-shaded areas denote the duration of the lockdown, while the green shaded area denotes the period when the test-and-isolate strategy is implemented. In the left-hand panel, each point represents a scenario drawn from the range of parameter values listed in Table 1; the spread of these points therefore captures the uncertainty in these different parameters.

More broadly, the ‘band’ of points in Figure 3 demarcates two distinct zones. Parameters in the lower zone correspond to testing strategies that prevent a resurgent epidemic from overwhelming the health system; and vice versa for parameters in the upper zone. These zones are illustrated by points P1 and P2, both corresponding to 20% seroprevalence at the point of lifting the lockdown, but with two different levels of testing effort (see Figure caption for details). As illustrated by the dynamics on the right-hand side, a resurgent epidemic overwhelms health system capacity in scenario P1 but not in P2.

Figure 4 shows these results under different scenarios for the basic reproduction number, *R*_0_. The dark-coloured points in both panels correspond to a testing strategy targeting solely symptomatic individuals (thus the dark blue points are identical to those shown in Figure 3). A comparison of dark-coloured points across the two panels illustrates that, for a given level of seroprevalence in the population, more intensive testing is needed at higher levels of *R*_0_, to prevent a resurgent epidemic from overwhelming hospital bed capacity. Taking the example of *R*_0_ = 3 (right-hand panel), at a population immunity of 20%, symptomatic testing would need to identify cases within 2.5 – 4 days on symptom onset. Such speed is unlikely to be infeasible in practice; in these situations it would be necessary to expand the testing strategy to include asymptomatic infection. The light-coloured dots in both panels show results for a testing programme that combines symptomatic testing with additional efforts to identify and isolate half of asymptomatic infections before they develop symptoms or recover. Once again, the y-axis shows the timeliness that is required, of the symptomatic-focused arm of this combined strategy. As would be expected, the inclusion of asymptomatic infections in the testing strategy has the effect of reducing the required intensity of symptomatic testing. Taking once again the example of 20% seroprevalence with *R*_0_ = 3, the symptomatic testing arm of a combined strategy would need to identify symptomatic infections between 7–8 days of symptom onset. We note that these results do not address how such testing performance might be met in practice, only the potential epidemiological implications of doing so. Below we briefly discuss implications for implementation.

**Figure 4.**
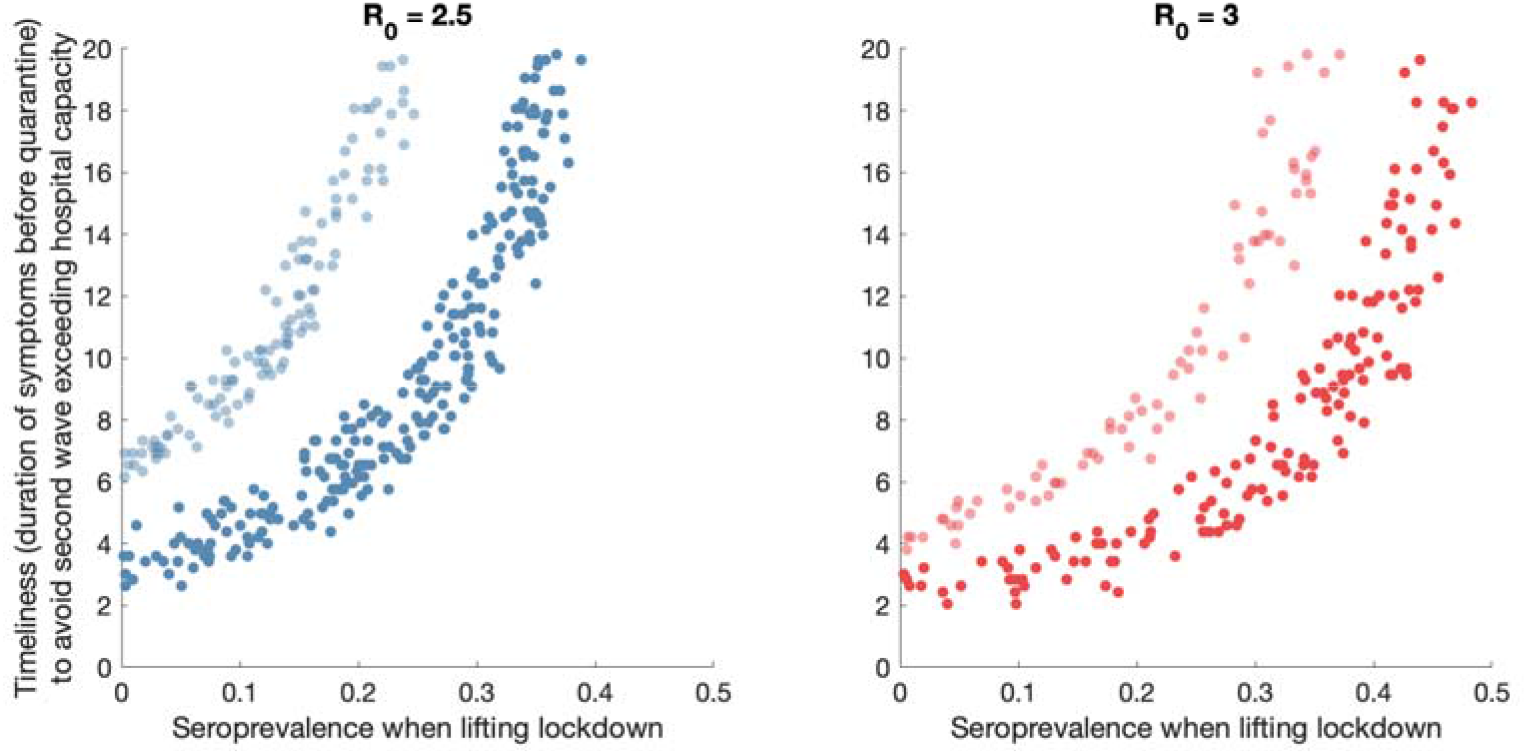
Sensitivity analysis to R0, and to testing strategy. Graphs are constructed as in Figure 3, left-hand panel. Figure shows results for R0 = 2.5 (left-hand panel) and for R0 = 3 (right-hand panel). The band of dark-shaded points corresponds to a strategy where only symptomatic cases are tested and isolated. The band of light-shaded points corresponds to a strategy where 50% of asymptomatic cases are additionally tested and isolated, before they recover or develop symptoms. As in Figure 3, the vertical axis denotes the timeliness with which symptomatic cases need to be identified and isolated, in order to prevent a resurgent epidemic from overwhelming the health system.

In the economic analysis to set these findings in context, we calculated the productivity loss in the megacity due to lockdown to be between 58% to 83.4%, translating into an economic loss of $1372 – $1979 million per week. By comparison, for the PCR-based testing programme, we estimated the cost of each RT-PCR test to be between $34.1 – $53.8. Thus we estimate that a hypothetical, symptom-based testing programme that identifies cases within 5 days of symptom onset would cost $26.68 – 42.12 million per week. This estimate is an upper bound as we used an artificially resource-intensive scenario for testing. For reasons discussed below, testing costs in practice are likely to be substantially lower.

## Discussion

Accurate and timely data will be critical in strategic planning for the control of the SARSCoV-2 pandemic. In the early days of the pandemic, India was amongst several countries (including the USA) facing an acute shortage of testing capacity, rendering it infeasible to perform intensive testing at the community level^16^. Nonetheless, the current ‘lockdown’ offers an opportunity to build up this testing capacity to the levels required. Our analysis illustrates how – even if there is uncertainty about how well a lockdown is working – ongoing sero-surveillance can provide invaluable information about the ‘here and now’. In particular, it provides an estimate of the fraction of the population that could still support transmission if the lockdown were lifted. In turn, the information from sero-surveillance can inform the intensity of effort that is needed, by any RT-PCR-based test-and-isolate strategy that aims to suppress transmission (Figures 3 and 4).

For the purpose of costing, we have adopted a simple, hypothetical scenario of symptom-based testing. As cautioned above, in practice it is unrealistic to test every individual showing COVID-19-like symptoms, particularly in settings such as Indian megacities (below we discuss alternative, more feasible approaches for timely case detection). Nonetheless, this scenario serves as a helpful upper bound, for the potential cost of a testing-based campaign. Our estimates suggest that even such a resource-intensive strategy would cost the government exchequer less than 2.1% of the projected economic loss. As another benchmark, this corresponds to less than half of the lockdown-caused loss in goods and services tax (GST) revenue^17^.

Our results highlight the importance of identifying and isolating cases as rapidly as possible, but do not capture ways in which this speed of response could be achieved. Contact tracing is one possible approach that is already being implemented in parts of India having a well-organised public health cadre, for example Kerala^1^. Recent evidence from China suggests that contact tracing could reduce the delay from symptom onset to isolation by 2 days^18^. This strategy is best suited to low-prevalence conditions, where (i) the volume of contacts to be investigated are within levels manageable by the health system, and (ii) the majority of infections are amongst contacts of known cases. Therefore, contact tracing could have strong value in the kind of testing programme that we have modelled here, with the purpose of maintaining low prevalence while allowing a lockdown to be lifted. Additionally, approaches such as pooling samples (while allowing for potential losses in accuracy) can help to use limited capacity to increase population coverage of testing^19,20^. Combinations of these strategies could also be implemented, for example, in the scenario of household contact tracing, with all samples from a household being pooled, and with all members of the household being quarantined if the pooled sample tests positive.

Modelling such strategies is outside the scope of our current analysis, but an important area for future work. We also note that our work does not address the performance requirements, of the different types of test that we model. For antibody tests in particular, there is wide recognition of the potential risks arising from imperfect specificity, at the individual level, i.e., that false positive results mean false assurance of immune protection^9,21^. In the present work, we only consider the use of such tests to assess immunity at the *population* level, where it may be possible to adjust for imperfect sensitivity and specificity (upto a certain level), in estimating seroprevalence^22,23^. In doing so, ethical considerations may require that participants in the test cohort remain uninformed of the results. We have also assumed that the testing for such surveillance will be conducted using a laboratory based test, where test accuracy is superior to point-of-care or home-based tests^24^.

At the time of writing, only a few surveys of seroprevalence to SARS-CoV-2 have been conducted around the world, with studies in the USA suggesting a seroprevalence of around 2 – 4% in different communities^25^. There remains intensive discussion about the validity of these and other early seroprevalence surveys^26^, but lessons learned from these approaches will be invaluable in other country settings. The use of well-validated antibody tests will be critical, as well as ensuring a representative population. The testing for such surveillance will need to be conducted at the laboratory level, where test accuracy is superior to point-of-care or home-based tests^24^. More broadly, in densely populated megacities with significant slum populations such as New Delhi or Mumbai, it is plausible that – despite stringent control measures during the lockdown – the proportion of the population ultimately exposed to the virus would reach higher levels than in settings such as the USA. Example scenarios, of 10 – 20% seroprevalence considered in Figure 3, may therefore not be implausible in Indian megacities, by the time the lockdown starts being lifted. At the time of writing, no such survey has yet been completed in Asia or in other LMIC settings, but the Indian Council of Medical Research has initiated a population-based sero-survey among adults in India^27^.

As with any modelling study, our approach has several limitations to note. As noted above, it offers illustrative scenarios using simple models of transmission. More detailed models could be helpful in planning the implementation of actual testing strategies. Such models could take account of population structure, including the elevated levels of transmission that might be expected in areas of high crowding, as well as incorporating household contact structure in a more explicit way than has been possible here. Fine-grained surveillance data would be invaluable in adequately parameterising such models. Such data might also allow the lifting of the lockdown to performed at a more granular geographic scale than what we have modelled here, for example if certain zones within a city show higher rates of infection than others. Our model currently does not accommodate these strategies, but could be modified accordingly. As illustrated by Figures 3 and 4, the approach that we propose – that of combining sero-surveillance and RT-PCR-testing – is also dependent on knowledge of the basic reproduction number, R0. Once again, robust surveillance data – even at the level of hospitalisations and mortality – can offer invaluable information for estimating this parameter over time^4^. Systematic follow-up of a cohort of cases and their contacts can also provide helpful data in this regard. We have only considered the role of testing and isolation in lifting a lockdown, whereas in practice this strategy might be combined with other measures, including maintaining physical distancing in shopping areas, public transport and other congregate settings. Depending on their effectiveness in reducing community transmission, such measures would be expected to shift the curves shown in Figure 3 upwards: that is, lowering the intensity that is required, of test-and-isolate programmes. Further data on the effectiveness of these interventions will be invaluable in refining our model findings accordingly. Our model is subject to various sources of uncertainty as well. Although we have incorporated wide uncertainty in lockdown effectiveness, as well as in the role of asymptomatic infection for transmission, there remain unanswered questions such as the role of children in transmission^28,29^, and the extent to which severity estimates, drawn from other settings^13^, may apply to Indian populations.

On the costing side, although our cost estimate is lower than the current ceiling price of $65 suggested by Indian Council of Medical Research for the private sector^30^, since we do not include the profit margins of the private sector, lower proportion of samples collected from home and increased scale of testing. In our estimates of economic cost of lockdown, we consider only productivity loss, which may be a reasonable approach in the immediate short term. However, the economic loss due to lockdown in the long run, is likely to be far greater as it would include demand side effects due to loss of employment and bankruptcies^31^.

Despite these caveats, we expect the fundamental point of our study to hold true: given the pressing need for evidence-based approaches towards lifting a lockdown, and in the face of ongoing epidemiological uncertainty, systematic sero-surveillance can be an invaluable source of evidence which can inform RT-PCR-based testing approaches in the community.

## Data Availability

The data generated or analysed during this study are included in this published article and its supplementary information file.

## Author contributions

NA and SD conceptualised the study. SM and HD conducted the analysis, and NA and SD validated the findings. SM and NA wrote a first draft of the manuscript, and all authors contributed to the final draft.

## References

1. Editorial. India under COVID-19 lockdown. Lancet 395, P1315 (2020).

2. Ministry of Health and Family Welfare.

3. Tian, H. et al. An investigation of transmission control measures during the first 50 days of the COVID-19 epidemic in China. Science (80-.). (2020). doi:10.1126/science.abb6105

4. Seth, F., Swapnil, M., Axel, G. & Al, E. Estimating the number of infections and the impact of non-pharmaceutical interventions on COVID-19 in 11 European countries. (2020). doi:https://doi.org/10.25561/77731

5. Marius, G., Mathias, D., Eric, M., Jean-Philippe, P. & Goldman, M. Preparing for a responsible lockdown exit strategy. Nat. Med. 1–2 (2020).

6. Ferguson, N. M., Laydon, D., Gemma, N.-G. & et al. Impact of non-pharmaceutical interventions (NPIs) to reduce COVID-19 mortality and healthcare demand. (2020). doi:https://doi.org/10.25561/77482

7. Prem, K. et al. The effect of control strategies to reduce social mixing on outcomes of the COVID-19 epidemic in Wuhan, China: a modelling study. Lancet Public Heal. (2020). doi:10.1016/s2468-2667(20)30073-6

8. Kissler, S. M., Tedijanto, C., Goldstein, E., Grad, Y. H. & Lipsitch, M. Projecting the transmission dynamics of SARS-CoV-2 through the post-pandemic period. medRxiv (2020). doi:10.1101/2020.03.04.20031112

9. World Health Organization. ‘Immunity passports’ in the context of COVID-19. (2020).

10. Financial Times. Indian coronavirus lockdown triggers exodus to rural areas. (2020).

11. Normile, D. Coronavirus cases have dropped sharply in South Korea. What’s the secret to its success? Science (80-.). (2020). doi:10.1126/science.abb7566

12. Anderson, R. M., Heesterbeek, H., Klinkenberg, D. & Hollingsworth, T. D. How will country-based mitigation measures influence the course of the COVID-19 epidemic? The Lancet (2020). doi:10.1016/S0140-736(20)30567-5

13. Verity, R. et al. Estimates of the severity of coronavirus disease 2019: a model-based analysis. Lancet Infect. Dis. (2020). doi:10.1016/s1473-3099(20)30243-7

14. Mizumoto, K., Kagaya, K., Zarebski, A. & Chowell, G. Estimating the asymptomatic proportion of coronavirus disease 2019 (COVID-19) cases on board the Diamond Princess cruise ship, Yokohama, Japan, 2020. Eurosurveillance (2020). doi:10.2807/1560-7917.ES.2020.25.10.2000180

15. Ferretti, L. et al. Quantifying SARS-CoV-2 transmission suggests epidemic control with digital contact tracing. Science (80-.). (2020). doi:10.1126/science.abb6936

16. Sharfstein, J. M., Becker, S. J. & Mello, M. M. Diagnostic Testing for the Novel Coronavirus. JAMA (2020). doi:10.1001/jama.2020.3864

17. Prasad, G. GST collections for April and May set to decline drastically. Livemint News Available at: https://www.livemint.com/news/india/gst-revenue-for-april-may-set-to-falldrastically-11588167297581.html.

18. Bi, Q. et al. Epidemiology and Transmission of COVID-19 in Shenzhen China: Analysis of 391 cases and 1,286 of their close contacts. medRxiv (2020). doi:10.1101/2020.03.03.20028423

19. Hogan, C. A., Sahoo, M. K. & Pinsky, B. A. Sample Pooling as a Strategy to Detect Community Transmission of SARS-CoV-2. JAMA – Journal of the American Medical Association (2020). doi:10.1001/jama.2020.5445

20. Deckert, A., Ba rnighausen, T. & Kyei, N. Pooled-sample analysis strategies for COVID-19 mass testing: a simulation study [Submitted]. Bull World Heal. Organ doi:http://dx.doi.org/10.2471/BLT.20.257188

21. Phelan, A. COVID-19 immunity passports and vaccination certificates: scientific, equitable, and legal challenges. Lancet (2020).

22. Diggle, P. J. Estimating Prevalence Using an Imperfect Test. Epidemiol. Res. Int. (2011). doi:10.1155/2011/608719

23. Lewis, F. I. & Torgerson, P. R. A tutorial in estimating the prevalence of disease in humans and animals in the absence of a gold standard diagnostic. Emerg. Themes Epidemiol. (2012). doi:10.1186/1742-7622-9-9

24. Adams, E. R. et al. Evaluation of antibody testing for SARS-Cov-2 using ELISA and lateral flow immunoassays. medRxiv (2020). doi:10.1101/2020.04.15.20066407

25. Bendavid, E. et al. COVID-19 Antibody Seroprevalence in Santa Clara County, California. medRxiv (2020). doi:10.1101/2020.04.14.20062463

26. Vogel, G. Antibody surveys suggesting vast undercount of coronavirus infections may be unreliable. Science (80-.). (2020). doi:10.1126/science.abc3831

27. Indian Council of Medical Research. National community based sero-survey for COVID-19. (2020).

28. Lee, P. I., Hu, Y. L., Chen, P. Y., Huang, Y. C. & Hsueh, P. R. Are children less susceptible to COVID-19? J. Microbiol. Immunol. Infect. (2020). doi:10.1016/j.jmii.2020.02.011

29. Qiu, H. et al. Clinical and epidemiological features of 36 children with coronavirus disease 2019 (COVID-19) in Zhejiang, China: an observational cohort study. Lancet Infect. Dis. (2020). doi:10.1016/S1473-3099(20)30198-5

30. Indian Council of Medical Research. Strategy of COVID19 Testing in India. (2020).

31. Bulsari, S. & Gumber, A. Economy’s Immunity against COVID-19. Econ. Polit. Wkly. 55, (2020).

32. Callow, K. A., Parry, H. F., Sergeant, M. & Tyrrell, D. A. J. The time course of the immune response to experimental coronavirus infection of man. Epidemiol. Infect. (1990). doi:10.1017/S0950268800048019

